# Spatial Correlation of Left Atrial Low Voltage Substrate in Sinus Rhythm versus Atrial Fibrillation: Identifying the Pathological Substrate Irrespective of the Rhythm

**DOI:** 10.1101/2022.02.18.22271172

**Authors:** Deborah Nairn, Martin Eichenlaub, Heiko Lehrmann, Björn Müller-Edenborn, Juan Chen, Taiyuan Huang, Claudia Nagel, Jorge Sánchez, Giorgio Luongo, Thomas Arentz, Olaf Dössel, Amir Jadidi, Axel Loewe

## Abstract

**Background:** Low voltage substrate (LVS) identified during electroanatomical mapping is a potential target for atrial fibrillation (AF) ablation. However, it is not clear how the location and extent of LVS correlate between mapping in sinus rhythm (SR) and AF.

**Objectives:** (1) Identify voltage dissimilarities between mapping in SR and AF. (2) Identification of regional voltage thresholds that improve cross-rhythm substrate detection. (3) Comparison of LVS between SR and native vs. induced AF.

**Methods:** Forty-one ablation-naive persistent AF patients underwent high-density voltage mapping in SR and AF. Each patient’s voltage information was mapped to a joint geometry for analysis. Global and regional voltage thresholds in AF were identified which best match LVS<0.5mV and <1.0mV in SR. Additionally, the correlation between SR-LVS with induced vs. native AF LVS was assessed.

**Results:** Substantial voltage differences (median: 0.52, IQR: 0.33-0.69, max: 1.19mV) with predominance of the posterior/inferior LA wall exist between the rhythms. An AF threshold of 0.34mV for the entire atrium provides an accuracy, sensitivity and specificity of 69%, 67% and 69% to identify SR-LVS<0.5mV, respectively. Lower thresholds for the posterior wall (0.27 mV) and inferior wall (0.3mV) result in higher spatial concordance to SR-LVS (4% and 7% increase). Concordance with SR-LVS was higher for induced AF compared to native AF (AUC: 0.80 vs. 0.73). AF-LVS<0.5mV corresponds to SR-LVS<0.97mV using high-definition mapping (AUC: 0.73).

**Conclusion:** The proposed voltage thresholds during AF maximise the consistency of LVS identification as determined during SR. Regional thresholds can further improve concordance.

## 1. Introduction

Atrial fibrillation (AF) is the most common sustained cardiac arrhythmia characterised by rapid irregular beating of the atria and associated with an increased risk for stroke and heart failure [1]. The pulmonary veins are the primary trigger site of AF. Isolating the pulmonary veins can yield a high rate of arrhythmia freedom in paroxysmal AF patients [2]. However, the success rate is often lower in persistent AF patients due to additional pathological substrate contributing to arrhythmia maintenance [3, 4, 5].

Electro-anatomical mapping to identify low bipolar voltages (peak-to-peak amplitudes) <0.5mV or 1mV during sinus rhythm (SR) have been shown to be a promising technique to identify the additional pathological substrate [4, 6, 7, 8, 9]. However, mapping in SR is not always feasible: e.g. when AF reoccurs shortly after electrical cardioversion due to recurrent / sustained fibrillatory trigger activity. Moreover, electrophysiologists may chose to perform mapping during AF, in order to identify both the potential arrhythmogenic rapid trigger sites and the underlying pathological substrate [10, 11, 12].

To date, two voltage cut-off values have been reported in SR that allow identification of potentially proarrhythmogenic tissue: <1mV in SR [13] and Rolf et al. reported ablation and isolation of atrial areas <0.5mV to improve SR maintenance rates at 12 months [6]. When mapping is done in AF, atrial areas displaying LVS <0.5mV have been reported as potential arrhythmogenic sites [4, 7]. Uncertainties remain regarding which cut-off values should be applied when mapping during AF and how the voltages in both rhythms relate to one another. The aim of the current study was to compare LVS in SR and AF and to identify regional voltage thresholds to improve cross-rhythm substrate detection.

## 2. Methods

### 2.1. Patient cohort

Forty-one patients with persistent AF presenting for their first AF ablation procedure were included in the study. Six weeks prior to AF ablation procedure, all patients were electrically cardioverted to SR, in order to enable favourable reverse electrical remodeling [14]. Patients presenting with SR on procedure date (11/41) underwent voltage and activation mapping in SR first, followed by voltage mapping during induced AF and subsequent PVI. Patients with native AF on procedure date (30/41) first underwent voltage mapping in AF, followed by electrical cardioversion, voltage and activation mapping in SR and finally PVI. Mapping was performed five minutes after AF induction, and only if AF was maintained for the entire mapping time. In two patients (5% of patients) with important sinus rhythm bradycardia and hypotension, pacing from the coronary sinus was performed.

### 2.2. Electro-anatomical Mapping

High-density voltage mapping was performed on forty-one persistent AF patients using the CARTO-3 mapping system (Biosense Webster, Diamond Bar, CA, USA) and a 20-pole (electrode size: 1 mm, spacing: 2–6–2 mm) Lasso-Nav catheter.

To avoid including information from points with poor contact, measurements were disregarded if the electrodes were located >6mm from the atrial surface. Band-pass filtering at 16-500 Hz was applied to the bipolar electrograms. To calculate the voltage, in SR a window of interest was chosen restricted to the PR interval in the ECG. In AF, the window of interest was set to include a single AF beat, with exclusion of QRS complex. Within this window, the peak-to-peak amplitude of the signal was calculated. Voltage values between the electrode positions were interpolated by the CARTO-3 system. Cut-off values of <0.5 and <1.0mV were then applied to the bipolar SR voltage maps to define the low voltage substrate (LVS) [4, 8]. Areas demonstrating LVS were confirmed using a separate contact force-sensing mapping catheter with a contact threshold of >5 g.

### 2.3. Analysis

Using the Scalismo statistical shape modelling software [15, 16, 17], the geometries of each patient were aligned and registered to a mean LA geometry [18]. This allowed the transfer of voltage information from each patient’s individual geometry to a common geometry represented by the same number of surface points, which represent the same anatomical landmarks. In this way, the analysis is not hindered by variations caused by spatial displacement. Additionally, the pulmonary veins and mitral valve areas could be easily treated separately during the analysis.

Equidistant points were chosen across the entire atria with a 3mm interpoint distance. The voltage value for each point was calculated as the mean amplitude of all points within a 1.5mm radius to compare local areas between the two rhythms.

To investigate the correlation between SR and AF, receiver operation curves (ROC) were created across the whole patient cohort, with the SR map being considered as the reference condition. The optimal AF thresholds for both SR cut-off values (0.5 and 1 mV) were identified and the sensitivity, specificity and accuracy were computed for each patient. In a subsequent local analysis, the percentage of patients showing consistent classification as low or high voltage in both rhythms per point was determined to identify the spatial concordance pattern.

A map of the median voltage values was constructed across the entire patient cohort for both rhythms. This allows a visual comparison between the maps without the influence of outliers due to patient-specific differences. Additionally, a map showing the difference between SR and AF median voltage values across all patients was computed.

The atrium was split into anatomical regions to examine the difference between SR and AF voltage mapping and identify optimal voltage thresholds for different atrial regions: inferior wall, lateral wall, posterior wall, anterior wall and roof. The optimal threshold for each region was identified as the top left-hand corner of the ROC curve when comparing the identification of low voltage regions between rhythms in that region.

Finally, the patient cohort was split into two groups: (1) where induced AF was mapped (patients presenting in SR) and (2) where native AF was mapped (patients presenting in AF). The two patients where the CS was paced to maintain SR were removed from this part of the study due to the unknown effect of how different fast pacing from the CS relates to AF. This group was too small to be considered separately. A two-sample t-test was performed to investigate if the voltage values between the two groups were significantly different. The two groups were then compared to their respective SR maps and ROC curves were computed. Since more patients were mapped with native AF (65%), a leave p-out (p=11) cross-validation was performed for the native ROC curve. A two-sample t-test was additionally performed between the distance of the two ROC curves in respect to the top left-hand corner.

## 3. Results

### 3.1. Patient characteristics

Forty-one persistent AF patients (63 ± 11.1 years old, 43.9% female) presenting for their first AF ablation procedure were included. On procedure date, 30 of 41 (73%) patients recurred with persistent AF. Table 1 describes further details regarding the patients’ characteristics.

**Table 1:** Patient clinical demographics. Abbreviations: BMI = body mass index, AA = antiarrhythmic, LA = left atrial, LVEF = left ventricular ejection fraction, LV = left ventricle, LVEDD = left-ventricular end-diastolic diameter

| **Patient Characteristics** | **Total = 41** |
| --- | --- |
| Age (years) | 63 ± 11.1 |
| Female (%) | 18 (43.9) |
| BMI (kg/sqm<sup>2</sup>) | 27.8 ± 6.1 |
| Arterial Hypertension (%) | 23 (56.1) |
| Diabetes mellitus (%) | 5 (12.2) |
| Prior stroke (%) | 1 (2.4) |
| structural CMP (%) | 12 (29.2) |
| Coronary artery disease (%) | 6 (14.6) |
| Persistent atrial fibrillation (%) | 41 (100) |
| CHA<sub>2</sub>DS<sub>2</sub>-VASc score | 1.7 ± 1.4 |
| Initial Rhythm SR (%) | 11 (26.8) |
| History of AF (months since diagnosis) (%) | 38 ± 41 |
| AA-therapy on admission (%) | 17 (41.5) |
| Betablocker Therapy Only (%) | 18 (43.9) |
| Amiodarone (%) | 11 (26.8) |
| Flecainide (%) | 3 (7.3) |
| Sotalol (%) | 1 (2.1) |
| Dronedarone (%) | 2 (4.8) |
| LA diameter (AP, mm) | 42.8 ± 6.9 |
| LVEF (%) | 50.4 ± 4.1 |
| LV dysfunction (LVEF<50%) | 6 (14.6) |
| LVEDD (mm) | 48.7 ± 6.1 |
| LA dilatation (>40 mm) | 28 ± 68.3 |
| Creatinine clearance (ml/min/1.73 sqm) | 74.1 ± 16.7 |

### 3.2. Spatial distribution of left atrial voltage during SR and AF

Figure 1 shows the distribution of LVS for a representative patient for mapping both during SR and AF. While the location of the LVS matches well between the rhythms on both the anterior and posterior wall, the extent is bigger in the AF map when using a cut-off value of 0.5mV for both maps. The voltage maps for all patients are shown in the supplementary material (Figure S1).

**Figure 1:**
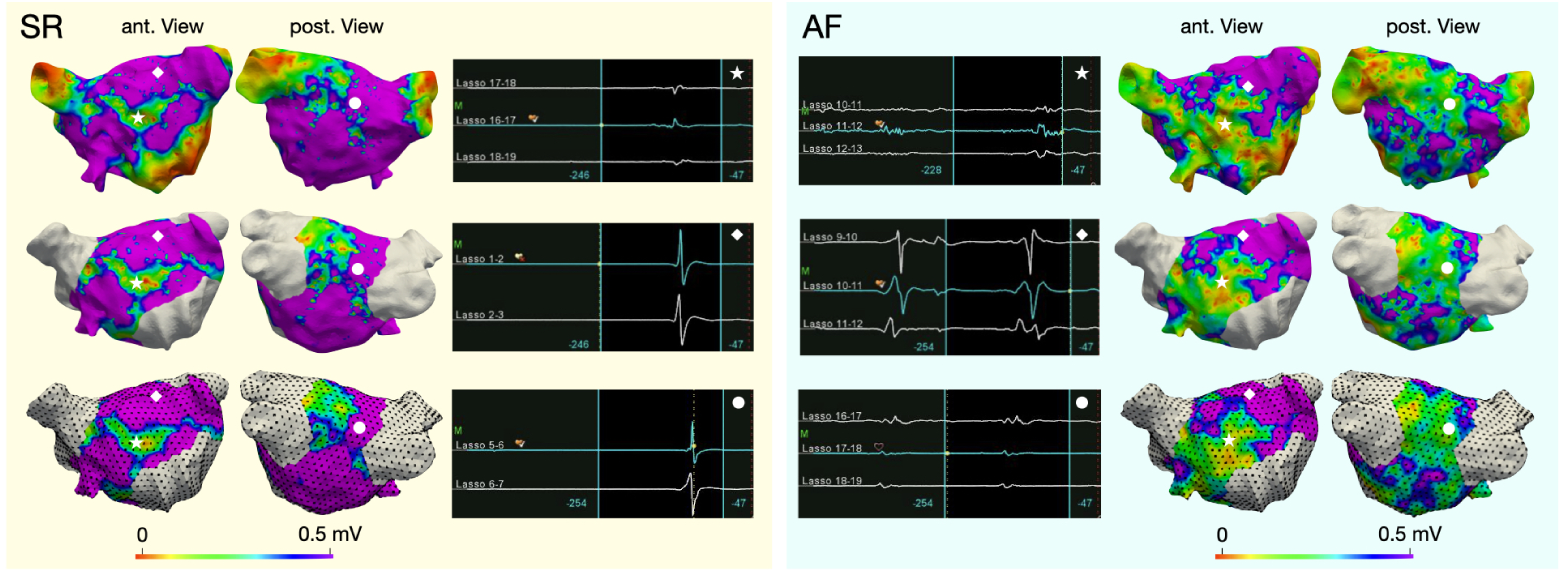
Spatial distribution of low voltage in SR and AF in one patient. Voltage maps in the top row are shown on the patient’s geometry. The middle row shows the voltage projected onto the joint geometry after exclusion of pulmonary vein and mitral valve areas. The bottom row shows the equidistant points (black dots) where the mean amplitude of all points within a 1.5 mm radius was considered. SR voltage map and relating signals are shown on the left (yellow box), AF on the right (blue box). The icons mark spatial points on the atrial geometry for which the corresponding signals are shown in the middle columns.

Figure 2 shows the spatial distribution of the median voltages across the entire patient cohort during both SR and AF. Voltages are lower across the entire atrium in AF (0.48±0.32 mV) than in SR (0.93±0.40 mV). The biggest differences between the median voltages in SR and AF of up to 1.1 mV are seen on the posterior and inferior wall. On the anterior wall, differences are smaller (0.35±0.14 mV). In both maps, the lowest voltages occur around the PVs and on the anterior wall (mean±std: 0.77±0.19 mV), with the highest voltages in the LAA (1.87±0.27 mV).

**Figure 2:**
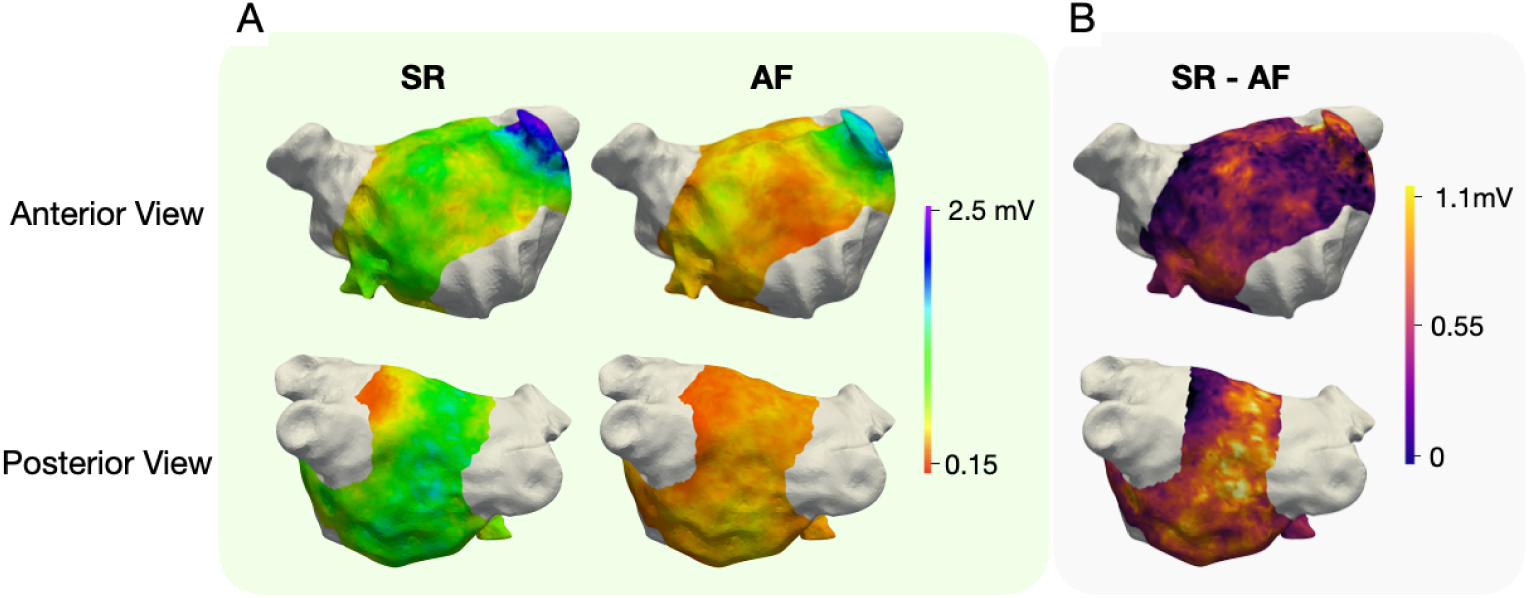
(A) Median voltage across all patients during SR and AF and (B) the difference of median voltage between the two rhythms.

### 3.3. Global AF thresholds for the detection of SR-LVS

ROC curve analysis provided the optimal threshold in AF for identifying LVS in SR. AF thresholds 0.34 and 0.45mV for SR <0.5mV and <1mV provided the best balance between high sensitivity and specificity as identified by the top left-hand corner of the ROC curve (Figure 3A). The percentage of concordance was moderate with a sensitivity of 67% (<0.5 mV) and 66% (<1 mV), specificity 69% (<0.5 mV) and 68% (<1 mV) and accuracy 69% (<0.5 mV) and 65% (<1 mV). The performance of the new AF voltage thresholds varied between patients with per-patient accuracy ranging between 53% and 94% (mean 69±11% for SR <0.5mV, mean 67±11% for SR <1 mV) (Figure 3B). In the supplementary material figure S2, the ROC curve identifying the optimal threshold in SR for AF <0.5mV is shown.

**Figure 3:**
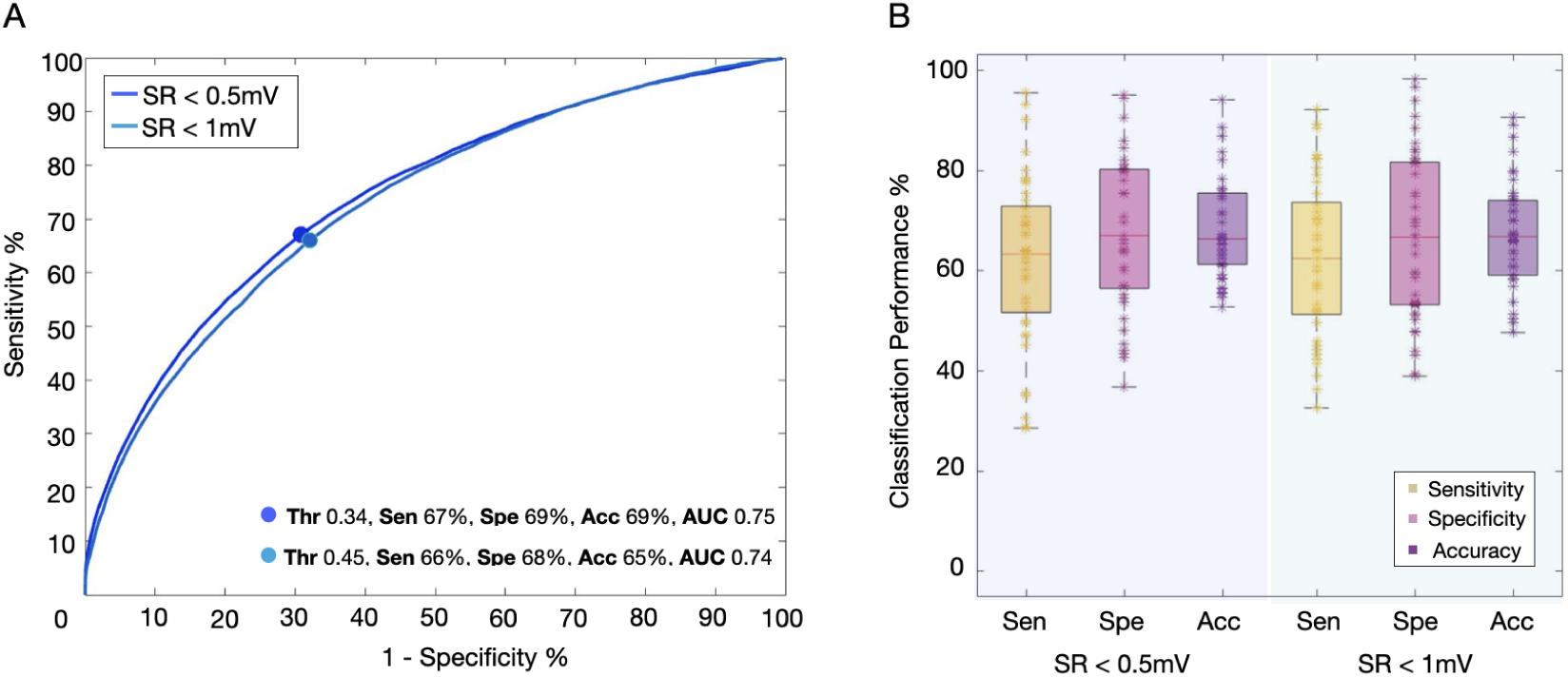
Cross-rhythm classification consistency. (A) ROC curve comparing LVS as identified during SR for bipolar thresholds <0.5mV and 1mV and during AF. The optimal AF thresholds across all patients and corresponding performance metrics are given in the legend. (B) classification performance parameters for each patient using the threshold obtained from the ROC curve.

Figure 4 shows the spatial distribution of the percentage of patients who demonstrate agreement of LVS areas in both SR and AF in a local neighbourhood. Agreement was high in the LAA (88±3% of patients). When using 0.5mV as the cut-off value in both SR and AF, there is agreement on the anterior wall (62±8%). However, the agreement is substantially smaller on the inferior and posterior walls (48±8%). By lowering the AF voltage thresholds to the values identified by the ROC curve analysis (figure 3A), the agreement on the anterior wall and inferior increased (69±8% and 61±8%).

**Figure 4:**
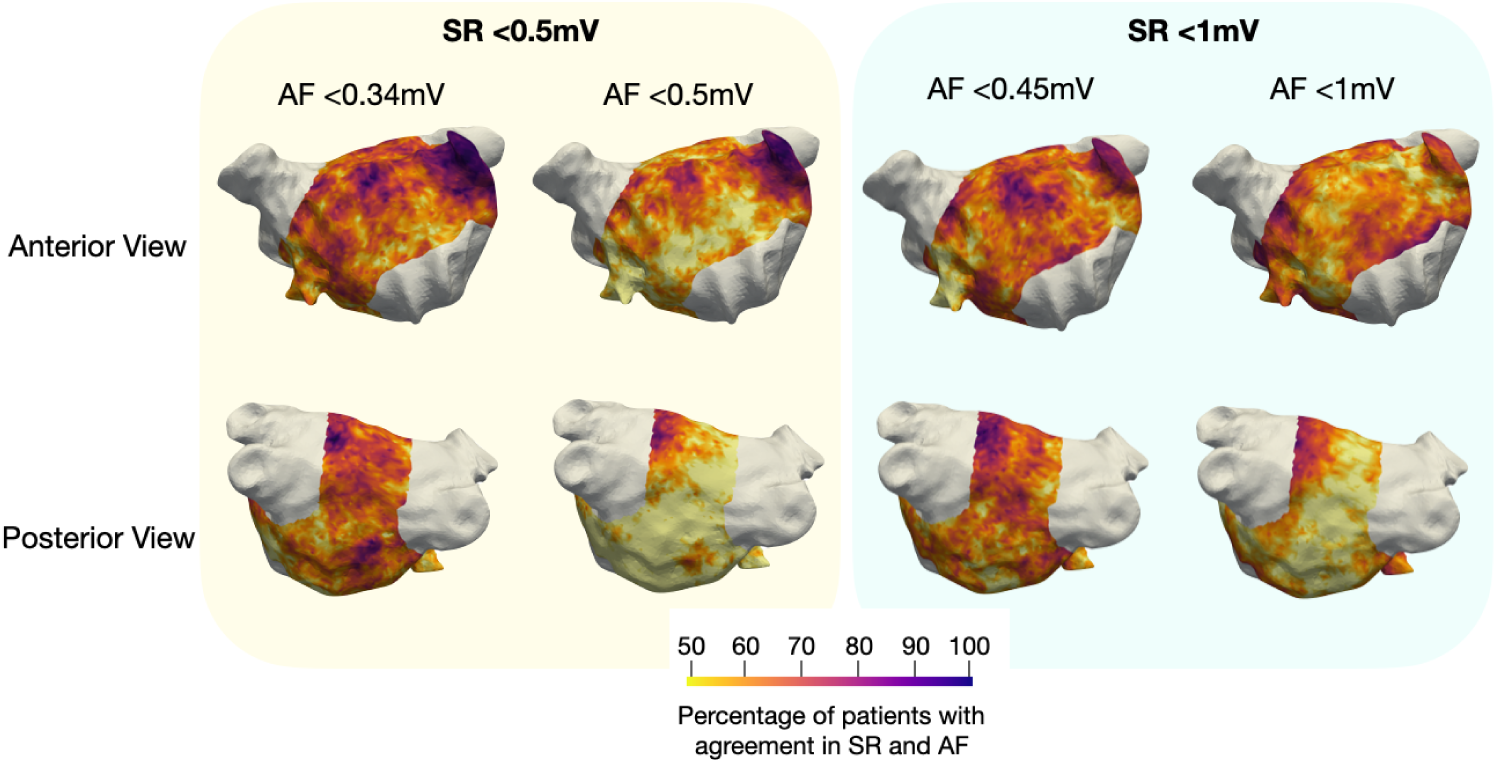
Agreement map between SR and AF. Each map shows the percentage of patients in which both voltage maps in SR and AF agree that low or high voltage is located within a local neighbourhood. The yellow box shows the agreement with SR <0.5mV, the light blue box with SR <1mV. In each box, the left column uses the optimal AF voltage thresholds as identified by the ROC curve analysis (Figure 3A).

**Figure 5:**
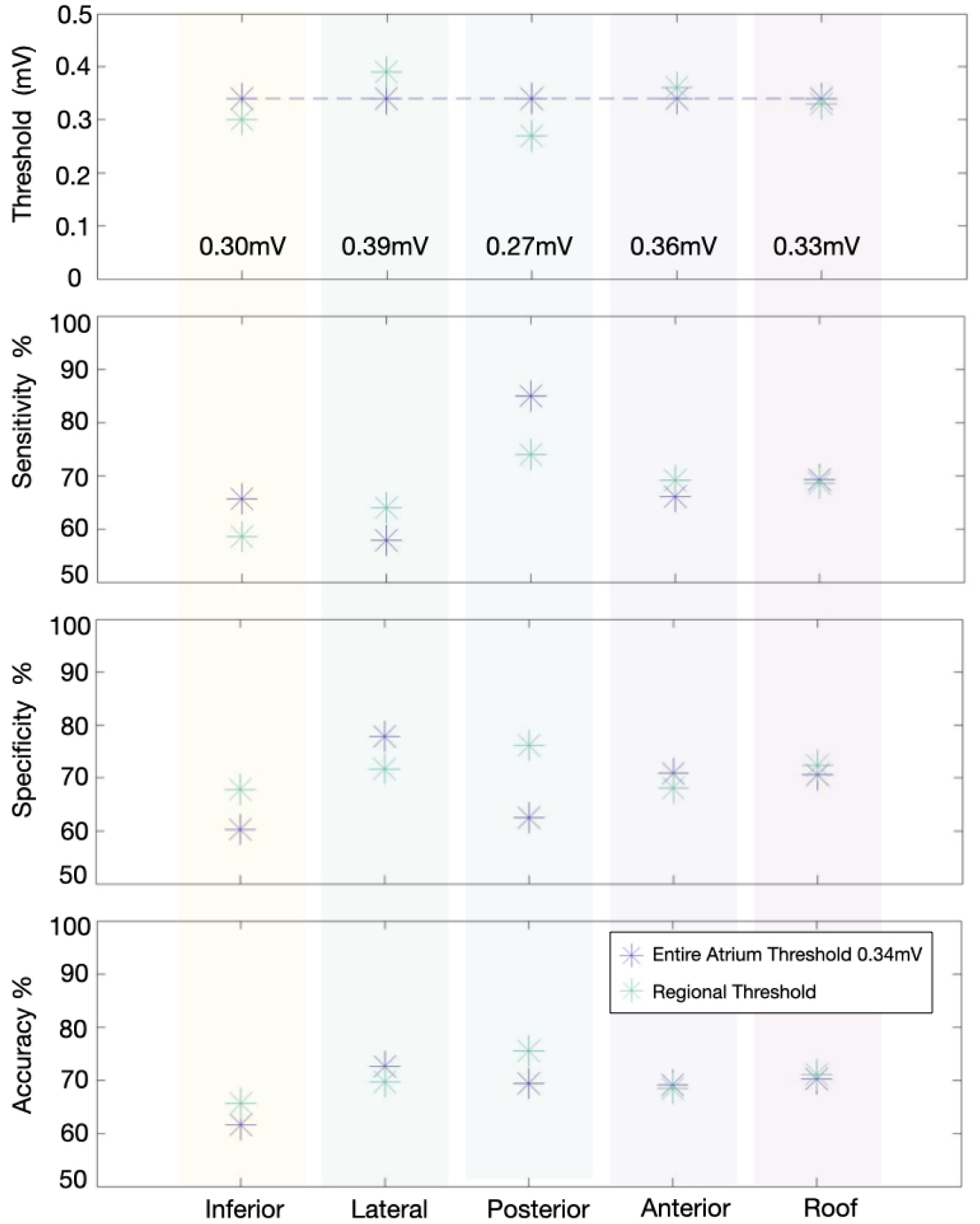
Optimal AF threshold and the corresponding sensitivity, specificity and accuracy for each anatomical region of the LA comparing to SR <0.5mV. The previously defined global threshold for the entire atria is shown by the purple asterisk while the green asterisk represents the regional threshold.

### 3.4. Regional AF thresholds for the detection of SR-LVS

The lowest concordance between SR and AF using the global optimal threshold for the entire atrium was found at the inferior wall (accuracy: 62%). By applying a regional AF threshold (0.3 mV), the accuracy on the inferior wall increased by 4%. For the posterior wall, an even lower AF threshold (0.27 mV) increased the accuracy from 69% to 76%. In both regions, the new regional thresholds decreased the sensitivity and increased the specificity. From figure 2 it can be seen that the voltage values are markedly higher on the posterior/inferior wall in SR than in AF. The entire atrium threshold is therefore too sensitive for these regions. On the other hand, on the anterior and lateral wall, a slightly higher threshold (0.36 and 0.39mV respectively) can optimally locate the regions of SR-LVS using the AF voltage map. The optimal AF regional thresholds which correspond to SR <1mV are shown in the supplementary material (Figure S3). The ROC curves used to find the optimal regional thresholds are shown in Figure S4.

### 3.5. Impact of inducing AF

The voltage was slightly higher in native AF patients than in patients in whom AF was induced (Figure 6A, not significant). The ROC curves (Figure 6B) show that the correlation between AF-LVS and SR-LVS is significantly (p<0.05) better in patients in whom AF was induced (AUC: 0.80 vs. 0.73). The optimal AF threshold for identifying SR-LVS <0.5mV was lower for patients in whom AF was induced (0.3 mV). In the supplementary material Figure S5, the boxplots and histograms can be seen for SR voltage values for patients who were cardioverted after mapping native AF versus first mapping native SR and then inducing AF. Although the difference was also not significant, the voltage values were typically higher in the patients mapped first in SR (median 1.66mV vs. 0.77 mV).

**Figure 6:**
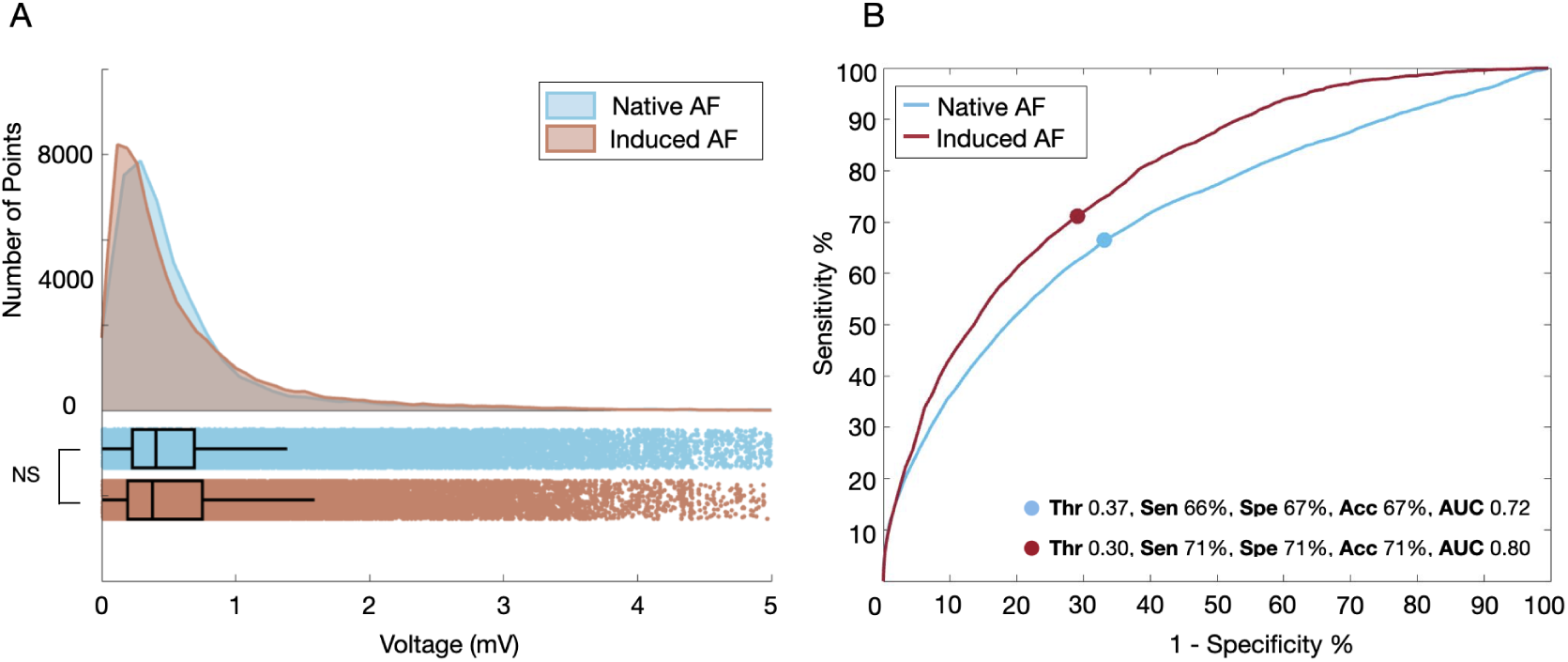
Histogram and boxplot of the voltage distribution in patients in whom AF was induced versus native AF. ROC curves comparing the AF-LVS in each group to SR-LVS. Blue: patients which presented with native AF, red: patients with induced AF.

## 4. Discussion

### 4.1. Main findings

This study investigated the differences in low voltage substrate identification for mapping during SR and AF. Three key findings can be reported:

1. The overall correspondence of LVS mapped in SR and in AF is moderate.
2. Discrepancies exist between mapping in SR and AF, specifically on the posterior and inferior LA wall.
3. New regional AF cut-off values improve detection of LVS as identified by mapping in SR.
4. The concordance of SR and AF voltage maps is higher when AF was induced compared to native AF.

### 4.2. AF cut-off values for identifying low voltage substrate

While cut-off values of <0.5 and 1mV are quite well established for mapping in SR [4, 8, 9], there is still no clear consensus whether the threshold applied when mapping during AF should be the same. A recent study using generalised additive models with a patient cohort of 31 patients found that a cut-off value of 0.31mV was best for predicting <0.5mV SR-LVS [19]. However, a point-by-point analysis was performed in this study, which could have been affected by undetected map shifts. To counter this problem, we analyse the data in small equidistant regions with a radius of 3 mm. Regardless, a similar cut-off value was found based on ROC curve analysis and local neighbourhood analysis (as opposed to point-by-point analysis) to determine the best cut-off value (0.34 mV). When examining LVS in AF as identified by regions <1mV in SR, a cut-off value of 0.45mV is optimal. However, as identified by the difference of the median voltage maps (figure 2), pronounced differences between the rhythms are present on the posterior and inferior walls. By adjusting the threshold for each region, the extent and location of SR-LVS can be better estimated when mapping during AF.

### 4.3. Differences in SR and AF voltage mapping

By examining the median voltages across the entire patient cohort for both rhythms, we showed that the voltages in SR are higher than in AF. This result is consistent with other studies [19, 7, 20] and can be due to multiple wave-fronts coming from various directions and endo-epicardial dissociation during AF [21]. Therefore, there are fewer coherent sources contributing to the electrograms. Moreover, neighbouring tissue may be depolarised by slightly delayed wavefronts, resulting in more fractionated signals and lower voltage amplitudes in AF [22]. This is further verified by the study from Ndrepepa el al., who identified that a reduction in voltage strongly correlates with the degree of AF disorganization [20].

While the voltage is typically lower in AF than SR, this study identified that this difference is not uniform across the entire atrium. The differences between rhythms were found to be much higher on the posterior and inferior LA wall (difference typically >0.55 mV) than on the anterior (difference typically <0.55 mV). Kurata et al. also reported higher voltages on the posterior region than the anterior region in both patients with and without low voltage areas [23]. Additionally, a recent study comparing voltage maps to LGE-MRI identified that the correlation between the two modalities was significantly better on the posterior wall when the voltage map was acquired during AF vs. SR [24]. One explanation drawn is that non-transmural or patchy fibrosis might be underestimated when mapping in SR. Sánchez et al. reported that low density fibrosis or electrode not located directly on top of the core of the fibrosis may cause discrete changes to the electrogram characteristics [25, 26]. On the other hand, when activation rates are more rapid during AF, non-transmural or patchy fibrotic tissue may be more susceptible to functional reentry, slow conduction or conduction block, which results in low voltage areas [27, 24]. However, catheter ablation of LGE-areas (including those at the posterior LA) did not improve the SR maintenance rates compared to PVI-only, in the large prospective multicenter trial DECAAF II [28]. Further computational studies may elucidate the mechanisms of rhythm-dependent voltage discrepancies on the posterior wall. Moreover, besides low voltage areas and LGE-areas, additional markers for arrhythmogenesis (e.g. repetitive rapid activity in AF or atrial late potentials in SR [12]) need to be considered by electrophysiologists to enable both a sensitive and specific detection of trigger sites for AF.

### 4.4. Influence of inducing AF in patients

We found that the correlation between SR and AF voltage mapping was better when the AF was induced in the patient. One hypothesis is that native AF is on average more complex, with high level of electrical remodeling, endo-epicardial dissociations of wavelet activities and more wavefronts originating from multiple directions than in induced AF that is mapped few minutes after its initiation [29, 21].

In the current study all persistent AF patients underwent electrical cardioversion to SR six weeks prior to scheduled AF ablation, in order to allow favourable reverse remodelling [14]. Patients who recurred with AF on the procedure date may have a more advanced form of electrical and structural LA remodelling than those who maintained SR. [30]. Thus, they may present with more complex patterns of propagation, such as multiple wavefronts and reentries. In this study, the mean voltages during SR are higher in the patients who presented with SR (median: 1.16mV, IQR: 0.51-2.18 mV) than those who recurred with AF (median: 0.77mV, IQR: 0.37-1.44 mV). A recent study reported that patients with extensive mean regional voltage reductions demonstrated whole LA degeneration [23]. This indicates a more advanced stage of arrhythmic remodelling in patients with AF recurrence six weeks after cardioversion to SR.

## 5. Limitations

The CARTO-3 mapping system was used for this study with a 20-pole Lasso catheter with electrodes of size 1 mm. Therefore, the AF thresholds may not apply to other systems or catheter configurations. In some patients, AF was mapped first and then the patient was cardioverted, potentially affecting the results by undetected map shifts. To counteract this, all patients’ voltage information was mapped to a joint geometry and analysis points comprised of the mean voltage in a 1.5 mm radius. The current study used high-density bipolar voltage mapping to compare LVS in SR and AF. The impact of omnipolar voltage mapping was not evaluated. However, comparing unipolar vs. bipolar voltage mapping modes, we recently reported that with high-density mapping, the direction of activation front to mapping electrodes does not significantly influence the extent and localization of detected LVS (both for SR and AF) [8].

## 6. Conclusion

The proposed AF thresholds improve the identification of SR-LVS, when mapping is performed during AF. However, a global threshold for the entire atria can lead to over-or underestimation of LVS, which can be corrected to some extent by applying the reported regional thresholds. Mapping in AF may be necessary in patients who cannot be cardioverted or maintained in SR or when AF mapping for detection of rapid activity sites is chosen. However, while application of the regional thresholds results in high concordances in LVS detection in about 70% of patients, significant discrepancies persist in the remaining patients. Further investigation needs to be undertaken to identify the role of complex propagation patterns in the characterisation of LVS when mapping during AF.

## Data Availability

The data analyzed in this study is subject to the following licenses/restrictions: To protect the safety of the patients, the data used for this study can not be provided. However, the figures within the article and Supplementary Material show detailed analyses for all patients used. Requests to access these datasets should be directed to: Deborah Nairn.

## Acknowledgements

We gratefully acknowledge financial support by Deutsche Forschungsgemein-schaft (DFG) through DO637/22-3, by the Ministerium für Wissenschaft, Forschung und Kunst Baden-Württemberg through the Research Seed Capital (RiSC) program.

## Notes

### Competing Interest Statement

The authors have declared no competing interest.

### Funding Statement

We gratefully acknowledge financial support by Deutsche Forschungsgemeinschaft (DFG) through DO637/22-3, by the Ministerium für Wissenschaft, Forschung und Kunst Baden-Württemberg through the Research Seed Capital (RiSC) program.

### Author Declarations

From 2017 to 2020, 41 patients with symptomatic persistent AF (lasting <7 days and <12 months) scheduled for their first PVI were included in this clinical study. The study was approved by the institutional ethics committee of the University of Freiburg (Germany) and all patients provided written informed consent.

