## Supplementary Material for "Spatial Correlation of Left Atrial Low Voltage Substrate in Sinus Rhythm versus Atrial Fibrillation: Identifying the Pathological Substrate Irrespective of the Rhythm"

### 1. Supplementary Figures

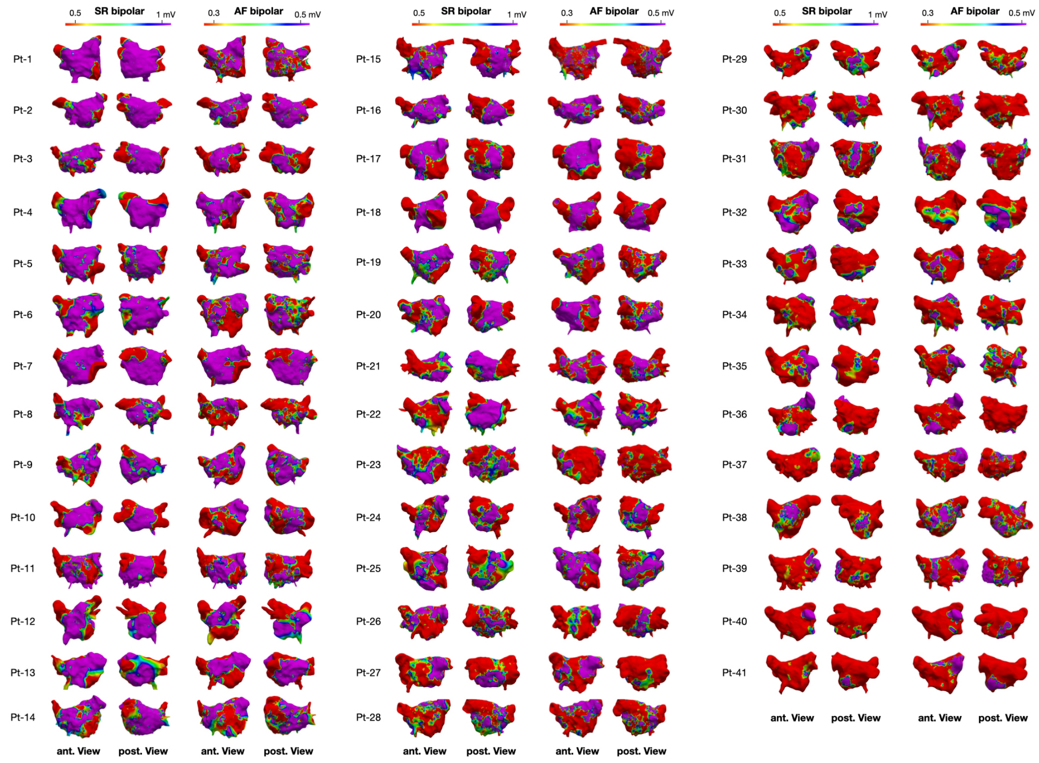


Figure 1: **Voltage maps acquired during SR and AF for all 41 patients.** For each patient the anterior (left) and posterior (right) view of the voltage map is shown. Cutoff values of 0.5-1 mV were set for the SR map and 0.3-0.5 mV for AF.

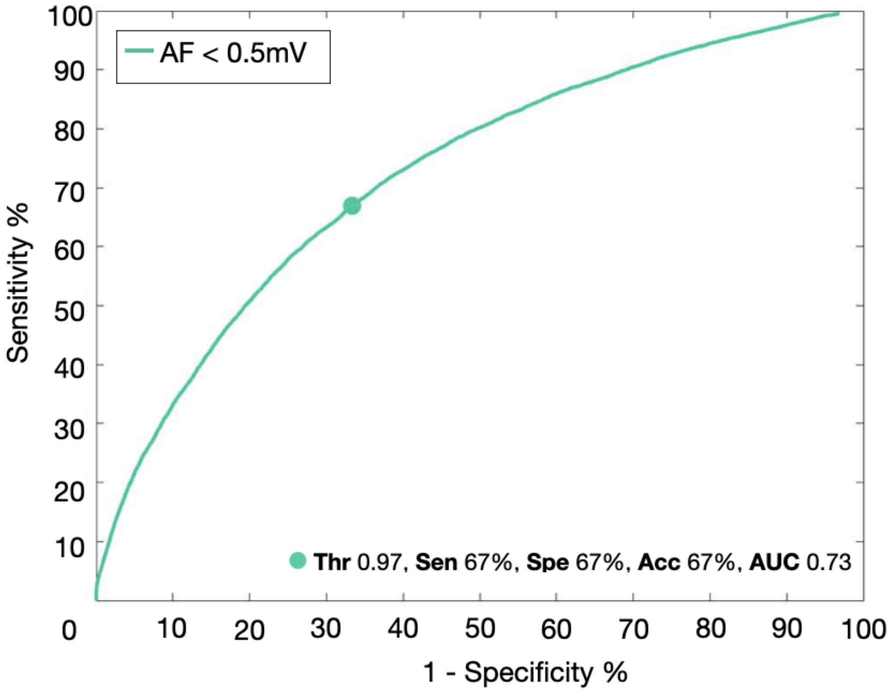


Figure 2: **ROC curve comparing LVS as identified during AF (<0.5 mV) and during SR .** The optimal SR threshold across all patients and corresponding performance metrics are given in the legend.

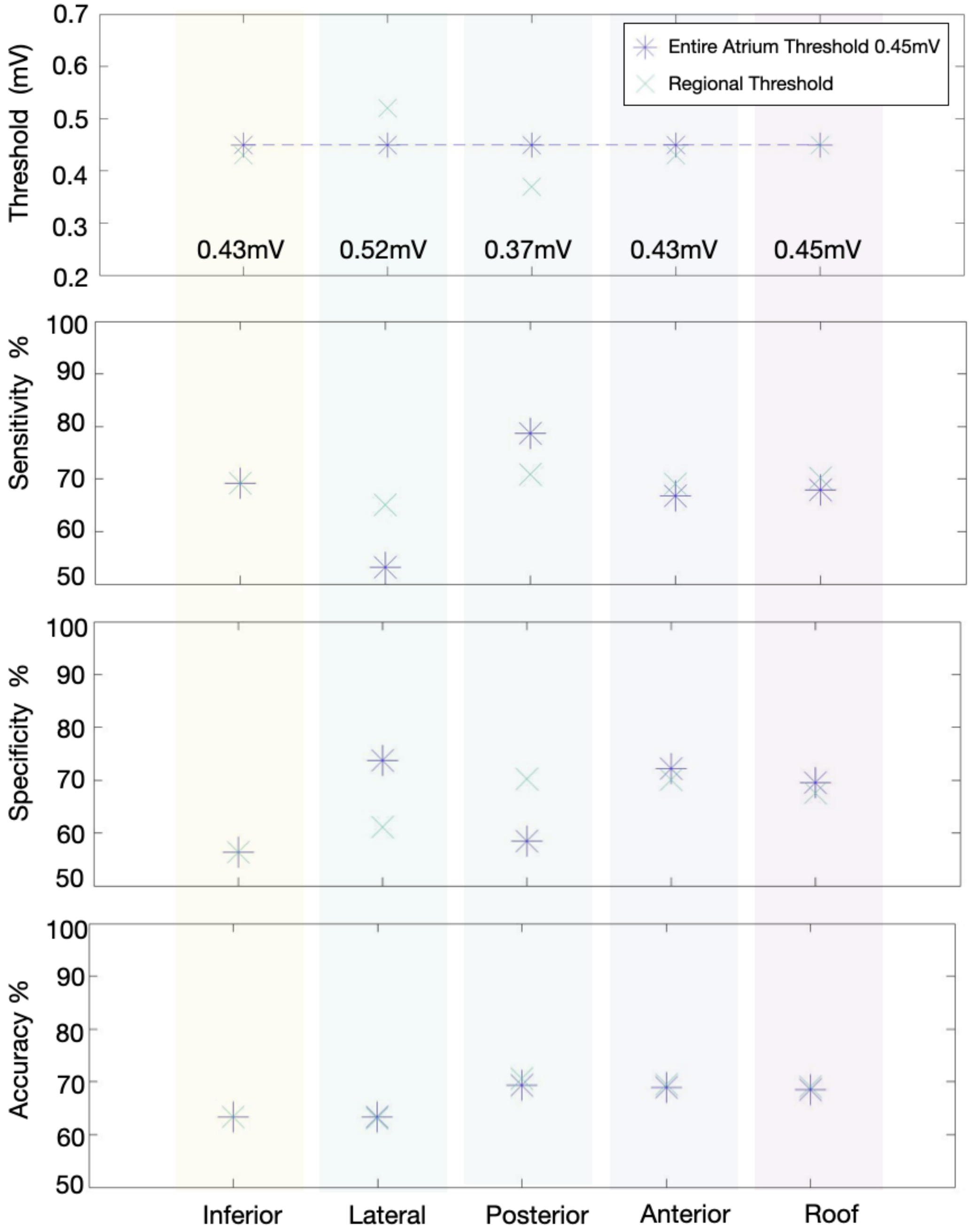


Figure 3: **Optimal AF threshold and the corresponding sensitivity, specificity and accuracy for each anatomical region of the LA comparing to SR-LVS <1 mV.** The previously defined global threshold for the entire atria is shown by the purple asterisk and the green cross presents the regional thresholds.

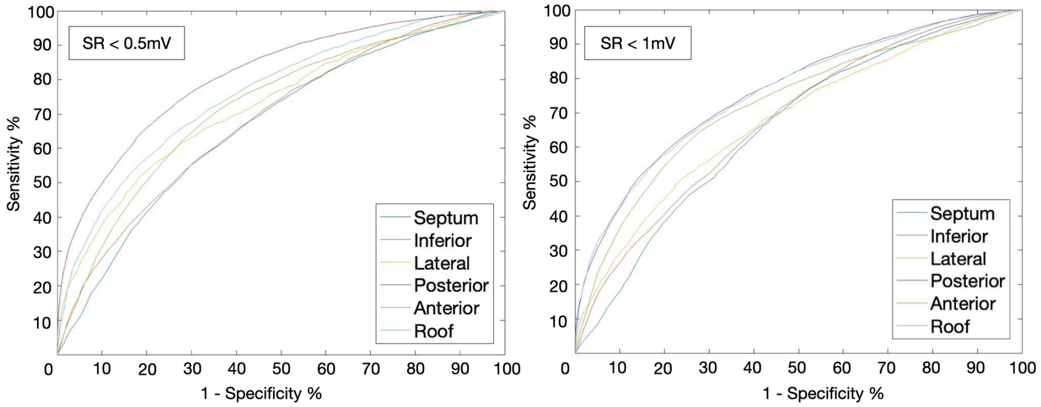


Figure 4: **ROC curves comparing the AF maps to the SR maps (<0.5 and <1 mV) for different anatomical regions of the left atria.**

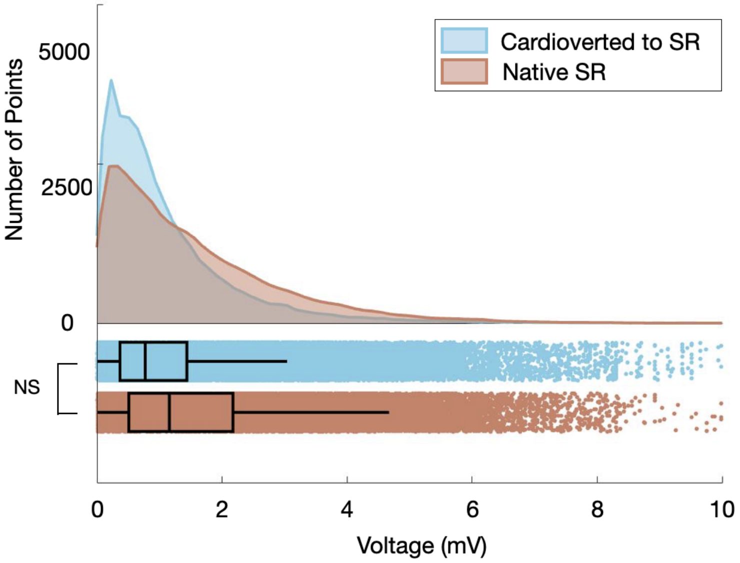


Figure 5: **Histogram and boxplot of the voltage distribution in patients in whom were cardioverted to SR (after mapping native AF) versus native SR.** Blue: patients whom were cardioverted to SR, red: patients presenting with native SR.
